# The efficacy of internet-based Cognitive Behavioral Therapy (CBT): Moodgym to help reduce depressive symptoms in repeating undergraduate students at The University of Zambia, Ridgeway campus

**DOI:** 10.64898/2026.02.12.26346135

**Authors:** Chimuka Muleya, Nasson Nathan Tembo, Steven Mukuka, Stanford Sakala, Taurai Maparuri, Himish Pual, Violet Muchimba, Joyce Ncheka, Ravi Paul

## Abstract

Depression is a common and clinically significant mental health condition among university students, particularly those experiencing academic failure and course repetition, and is associated with adverse effects on cognitive functioning, emotional regulation, and academic performance. This study evaluated the efficacy of an internet-based cognitive behavioural therapy (iCBT) intervention, MoodGYM, in reducing depressive symptoms among repeating undergraduate students at the University of Zambia Ridgeway Campus.

A quasi-experimental quantitative study design was employed. Seventy-five repeating undergraduate students with depressive symptoms were enrolled, with 33 assigned to the MoodGYM intervention group and 42 to a control group. Depressive symptom severity was assessed using the Beck Depression Inventory (BDI) at baseline and after an eight-week intervention period. Statistical analyses included within-group and between-group comparisons, difference-in-differences estimation, and fixed-effects regression modelling.

At baseline, participants exhibited predominantly moderate to severe depressive symptoms, with no statistically significant differences between the intervention and control groups. Following the eight-week intervention, the MoodGYM group demonstrated a statistically and clinically significant reduction in depressive symptoms, with median BDI scores decreasing from 22 to 16 (p < 0.001), representing a large effect size (Cohen’s d = 1.02). In contrast, the control group showed persistence or worsening of depressive symptoms over the same period. Difference-in-differences analysis confirmed a robust intervention effect, with an approximately 10-point greater reduction in depression scores among MoodGYM participants compared with controls (p < 0.001).

These findings indicate that MoodGYM is an effective internet-based intervention for reducing depressive symptoms among repeating undergraduate students and offers a feasible and scalable approach to addressing student mental health needs in low-resource university settings.

## Introduction

Depression is a leading cause of disability worldwide and constitutes a major public health concern, particularly among young adults in higher education (1,2). University students are exposed to multiple academic, social, and financial stressors that increase vulnerability to depressive symptoms, which in turn negatively affect cognitive functioning, emotional regulation, academic performance, and overall quality of life (3). Medical and health sciences students are especially susceptible due to intense academic demands, frequent examinations, competitive learning environments, and high personal expectations (4). Within this population, students who experience academic failure and are required to repeat courses face an additional psychological burden characterised by feelings of shame, hopelessness, reduced self-efficacy, and fear of further failure.

In sub-Saharan Africa, the burden of depression among university students is substantial, with reported prevalence estimates ranging from 23.3% to 76.5% (5–8). In Zambia, although direct prevalence data on depression among repeating undergraduate students are limited, evidence indicates extremely high levels of academic stress. A study conducted at the University of Zambia Ridgeway Campus reported academic pressure as the leading source of stress, affecting over 93% of undergraduate medical students, with the highest burden observed among third-year students—a cohort that includes a large proportion of repeating students (9). Academic failure and course repetition are well-established risk factors for depression, suggesting that repeating students constitute a particularly vulnerable and under-researched group.

Despite the high burden of depressive symptoms, access to mental-health care among university students remains poor. Studies consistently demonstrate low rates of help-seeking behaviour, driven by stigma, concerns about confidentiality, time constraints, cost, and fear of negative academic or professional consequences (10,11). Approximately 69% of students are unlikely to seek mental-health services, and those who perceive stigma are significantly less likely to access care (11). These barriers are particularly pronounced among repeating students, who are often expected to continue attending lectures and clinical placements while managing academic remediation, leaving little opportunity to attend regular face-to-face psychotherapy sessions (4).

Cognitive Behavioural Therapy (CBT) is widely recognised as an effective, evidence-based treatment for depression and is recommended as a first-line psychological intervention (12–14). However, in Zambia and many other low-income settings, the delivery of CBT is constrained by a severe shortage of trained mental-health professionals, under-resourced services, and limited national investment in mental health, which receives less than 1% of the health budget (15,16). These systemic constraints necessitate alternative, scalable approaches to delivering psychological care.

Internet-based psychological interventions, particularly internet-based cognitive behavioural therapy (iCBT), have emerged as a promising solution to address treatment gaps in mental-health care. iCBT programs adapt structured CBT principles into digital, self-guided formats that can be accessed privately, flexibly, and at low cost (17,18). Such interventions have demonstrated effectiveness in reducing symptoms of depression, anxiety, and stress across diverse populations, including university students (19,20).

One of the most extensively researched iCBT programs is MoodGYM, an interactive intervention developed by the National Institute for Mental Health Research. MoodGYM delivers CBT-based psychoeducation and skills training through five structured modules focusing on emotions, thoughts, cognitive distortions, stress management, and interpersonal relationships (21,22). Since its launch in 2001, MoodGYM has been evaluated across multiple countries, age groups, and settings, with evidence supporting its effectiveness in reducing depressive symptoms both with and without therapist guidance (23,24).

However, despite the growing global evidence base, the application of iCBT in low-resource African settings remains limited. In Zambia, only one prior study has examined MoodGYM among students, focusing primarily on feasibility during the COVID-19 pandemic, without a control group or quantitative assessment of efficacy (25). Furthermore, no published studies have specifically evaluated the effectiveness of iCBT interventions among repeating undergraduate students, a group at heightened risk of depression and academic disengagement.

The field of digital mental health is also characterised by ongoing debate. While proponents emphasise the scalability, cost-effectiveness, and stigma-reducing potential of iCBT, critics highlight limitations related to user engagement, cultural relevance, language complexity, and the absence of real-time therapeutic interaction (26,27). These debates underscore the importance of generating robust, context-specific quantitative evidence to inform the implementation of digital mental-health interventions.

Against this backdrop, the present study aimed to evaluate the efficacy of the internet-based cognitive behavioural therapy program MoodGYM in reducing depressive symptoms among repeating undergraduate students at the University of Zambia Ridgeway Campus. A quasi-experimental quantitative design was employed to compare changes in depressive symptom severity between students who received the MoodGYM intervention and a control group. The study demonstrated a statistically and clinically meaningful reduction in depressive symptoms among students who used MoodGYM compared with controls, thereby achieving its primary objective and providing locally relevant evidence to support the integration of internet-based cognitive behavioural therapy into university mental-health services in Zambia.

## Materials and Methods

### Study Design

A quasi-experimental quantitative study design was employed. This design was used to evaluate the effectiveness of the internet-based cognitive behavioural therapy (CBT) intervention, MoodGYM, in reducing depressive symptoms among repeating undergraduate students. The quasi-experimental approach was appropriate because random assignment of participants to intervention and control groups was not feasible within the academic setting. The design allowed for comparison of depressive symptom outcomes between students who received the MoodGYM intervention and those who did not, thereby enabling assessment of the intervention’s efficacy under real-world conditions.

### Sampling Technique

Stratified random sampling was used to select study participants. This technique ensured that key subgroups within the population were adequately represented by stratifying participants according to relevant characteristics such as year of study and gender. By doing so, the study enhanced the representativeness of the sample and improved the accuracy and generalisability of the findings among repeating undergraduate students.

### Study Site

The study was conducted at the University of Zambia Ridgeway Campus, located along the Nationalist Road in Lusaka, Zambia. This site was selected because it hosts the target population of repeating undergraduate students and provides an appropriate academic environment for implementing an internet-based intervention. Its central location and existing infrastructure supported efficient participant recruitment and delivery of the online intervention.

### Study Population

The study population comprised all repeating undergraduate students at the University of Zambia Ridgeway Campus. This group was considered appropriate for the study because repeating students face distinct academic pressures that increase their risk of depressive symptoms, making them an ideal population for evaluating the efficacy of the MoodGYM intervention.

### Sample size determination

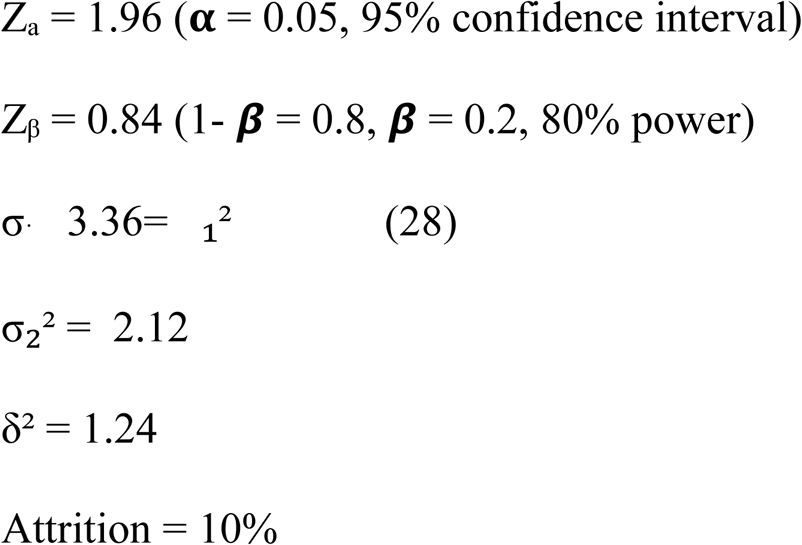

### Power Analysis Formula

This method is employed to calculate the necessary sample size for achieving statistical power. A power analysis ensures that the study has enough participants to d alletect a meaningful effect of MoodGym on depression symptoms, minimizing the risk of Type II errors. The formula takes into account expected effect size, significance level, and desired power, which are crucial for determining adequate sample size.

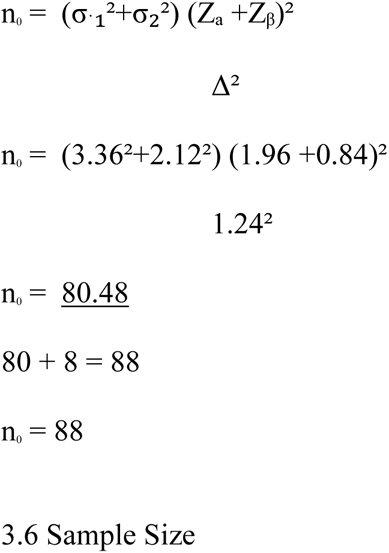

The calculated sample size for the quantitative component was 88 participants, based on power analysis to ensure adequate statistical power to detect a meaningful effect of the MoodGYM intervention in the study population. Although the target sample size was not fully achieved, a total of 75 participants were enrolled and analysed. This sample size remained statistically sufficient to allow for robust and meaningful quantitative analysis of the intervention’s effect.

### Selection Criteria

#### Inclusion Criteria

The study included repeating undergraduate students from the University of Zambia Ridgeway Campus across all programmes, including Medicine, Pharmacy, Physiotherapy, Nursing Sciences, and Environmental Health. Participants were required to be 18 years of age or older.

#### Exclusion Criteria

Students who were participating in another depression intervention study were excluded. In addition, students with depression secondary to other medical or psychiatric conditions were excluded, as determined through a brief medical history obtained prior to enrolment.

### Study Procedures by Specific Objective

#### Objective 1: To assess baseline levels of depressive symptoms among repeating undergraduate students

Baseline assessment of depressive symptoms was conducted using an online survey that collected demographic information and included the Beck Depression Inventory (BDI). The survey was distributed via Google Forms to class WhatsApp groups at the Ridgeway Campus after students had been fully informed about the study aims, objectives, risks, and benefits. From respondents, 178 repeating students were identified using stratified random sampling to ensure representation across year of study, academic programme, and severity of depressive symptoms. Students with BDI scores below 10 were excluded, as this indicated normal mood fluctuations, while those with BDI scores greater than 40 were referred to Clinic 6 at the University Teaching Hospital for pharmacological management (29,30). Written informed consent was obtained, and baseline BDI assessments were completed prior to implementation of the intervention.

#### Objective 2: To measure change in depressive symptoms following the MoodGYM intervention over eight weeks

Participants were allocated into two groups: an intervention group that received access to the MoodGYM program and a control group that did not receive any intervention during the study period. Participants in the control group were blinded to the intervention, and those in the intervention group were instructed not to share information about the program to minimise contamination. The MoodGYM group engaged with the internet-based intervention for eight weeks, with a minimum recommended usage of two sessions per week (21,23,24). Depressive symptoms were assessed using the BDI at baseline and again at week eight. Adherence was monitored through scheduled check-ins at weeks 1, 3, and 6, while intervention fidelity was verified using MoodGYM login data and module completion logs. An acceptable loss-to-follow-up rate of 20% was predefined (29), and intention-to-treat analysis was applied to manage incomplete data.

#### Objective 3: To compare depressive symptom outcomes between intervention and control groups

Both intervention and control groups completed BDI assessments at identical time points. Changes in depressive symptom scores between groups were compared using quantitative statistical methods, including difference-in-differences estimation, to determine the relative effectiveness of the MoodGYM intervention. Measures to reduce contamination included secure individual login credentials, single blinding of the control group, participant instructions to avoid information sharing, and separation of WhatsApp communication groups for intervention and control participants. A per-protocol analysis was also conducted to assess the intervention effect among participants who adhered to the study protocol.

### Control Group

The control group consisted of repeating undergraduate students who did not receive the MoodGYM intervention during the study period and served as a comparison group for evaluating the effect of the internet-based CBT program. Participants were selected using stratified random sampling from the initial pool of 178 eligible students to ensure balanced representation across key demographic and academic characteristics. Throughout the study, strict procedures were implemented to prevent contamination between groups, including separation of study communications and restricted access to intervention materials. Upon completion of the study, a post-study “flip-over” procedure was implemented, allowing participants in the control group to access the MoodGYM intervention, thereby ensuring ethical equity and potential mental health benefit for all participants.

### Data Analysis

**Table 1:** Data Analysis.

| Specific Objective | Type of Analysis | Type of Statistics |
| --- | --- | --- |
| To assess the baseline levels of depression symptoms in repeating undergraduate students at The University of Zambia Ridgeway Campus before the implementation of the Moodgym program | Mean, Mode, Medium, SD, Variance | Descriptive statistics |
| To measure the change in depression symptoms among these students after participating in the Moodgym program over a period 8 Weeks. | Difference of difference and Regression | Inferential statistics |
| To compare the depression symptom outcomes between students who participated in the Moodgym program and those who did not, to determine the relative effectiveness of this intervention. | Difference of difference and Regression | Inferential statistics |

**Table 1:** Study Variables.

| Independent Variable | Scale of Measure | Dependent Variable | Scale of Measure |
| --- | --- | --- | --- |
| Age | Continuous - Ordinal | BDI Score | Discrete – Ordinal |
| Sex | Categorical - Binary | Depression | Binary |
| Residence | Continuous - Nominal |  |  |
| Program of Study | Categorical - Nominal |  |  |
| Year of Study | Categorical - Ordinal |  |  |
| Number of course repeated | Continuous - Ordinal |  |  |
| Number of Times repeating | Continuous - Ordinal |  |  |

### Ethics Considerations

Following receipt of a clearance letter from the Assistant Dean of Postgraduate Studies, ethical approval for this study was obtained from the University of Zambia Biomedical Research Ethics Committee (UNZABREC), in accordance with institutional and national ethical requirements. Additional administrative permission to conduct the study was granted by the Assistant Deans of the Undergraduate Schools of Medicine, Nursing Sciences, Public Health, and Health Sciences.

Participation in the study was entirely voluntary. Written informed consent was obtained from all repeating undergraduate students prior to enrolment. The consent process involved a comprehensive explanation of the study objectives, procedures, potential risks and benefits, the voluntary nature of participation, and the right to withdraw from the study at any time without providing a reason and without any academic or personal consequences. Written consent was documented using a standardized informed consent form, which was signed and dated by each participant in the presence of an independent witness, who subsequently co-signed the form to confirm that the information had been adequately explained and understood. All signed consent forms were securely stored and maintained as part of the study records.

Data collection for the MoodGYM study was conducted over a six-month period, commencing on 01 February 2025 and concluding on 29 July 2025. During this period, participants were recruited and study questionnaires were administered in accordance with the approved study protocol. Participants allocated to the intervention group were required to engage with the MoodGYM program for approximately two hours per week over an eight-week period. The internet-based nature of the intervention allowed flexible participation, minimising disruption to academic schedules and reducing the burden associated with fixed, appointment-based psychotherapy.

Confidentiality was strictly maintained throughout the study. All data collected were used solely for research purposes, and personal identifiers were removed during data processing. Unique identification codes were assigned to each participant to ensure anonymity during data analysis and reporting.

The principal investigator assumed full responsibility for the conduct of the study and implemented all reasonable measures to minimise potential risks to participants. Study procedures were designed to reduce the likelihood of psychological distress, including clear communication regarding voluntary participation and the non-evaluative nature of the intervention. Informed consent was obtained without coercion, and participants retained full autonomy over their continued involvement in the study.

Given the sensitive nature of assessing depressive symptoms, additional safeguards were implemented to protect participant well-being. Participants who recorded very high depression scores or demonstrated worsening symptoms during the study were promptly referred to Clinic 6 at the University Teaching Hospital for further clinical evaluation and management. These measures ensured that participant safety and well-being remained a priority throughout the research process.

## Results

### Demographic Characteristics

#### Introduction

This chapter presents the quantitative findings on the effect of the internet-based cognitive behavioural therapy (CBT) program, MoodGYM, on depressive symptoms among repeating undergraduate students at the University of Zambia Ridgeway Campus. A total of 75 students participated in the study, comprising 42 students in the control group and 33 students in the intervention group. Data were cleaned and analysed using R version 4.5.2 for Windows (R Core Team, 2025). Descriptive statistics were summarised using frequencies and percentages for categorical variables, and means with standard deviations or medians with interquartile ranges for continuous variables, depending on data distribution.

Baseline comparability of depressive symptom scores and other independent variables between the intervention and control groups was assessed using chi-square or Fisher’s exact tests for categorical variables, and independent t-tests or Wilcoxon rank-sum tests for continuous variables. Within-group changes in depression scores from baseline to post-intervention were analysed using Wilcoxon signed-rank tests. Between-group comparisons of change scores were conducted using independent t-tests or Mann–Whitney U tests, with effect sizes reported using Cohen’s d.

To estimate the intervention effect over time, a difference-in-differences (DiD) analysis was performed using a fixed-effects panel regression model, with the interaction between treatment group and time serving as the DiD estimator. The fixed-effects approach controlled for time-invariant individual characteristics, while robust standard errors clustered at the participant level accounted for heteroskedasticity and within-participant correlation. Model estimates, robust standard errors, and 95% confidence intervals were reported.

Sensitivity analysis was conducted using an ordinary linear regression model with post-intervention depression scores as the dependent variable, and treatment group, baseline depression scores, and relevant covariates as predictors. Graphical presentations included line plots illustrating pre- and post-intervention trajectories for both within-group and between-group comparisons. All statistical tests were two-sided and conducted at a significance level of 0.05.

#### Baseline Characteristics

The baseline characteristics of the 75 study participants are summarised in Table 3. The majority of participants were female (76.0%, n = 57), and most identified as Christian (96.0%, n = 72). More than half of the participants resided in boarding houses (57.3%, n = 43), while 17.3% (n = 13) lived at home and 25.3% (n = 19) resided on the Ridgeway Campus. Most participants were enrolled in the School of Medicine (57.3%, n = 43), followed by the School of Health Sciences (28.0%, n = 21), the School of Public Health (10.7%, n = 8), and the School of Nursing Sciences (4.0%, n = 3).

**Table 3:**
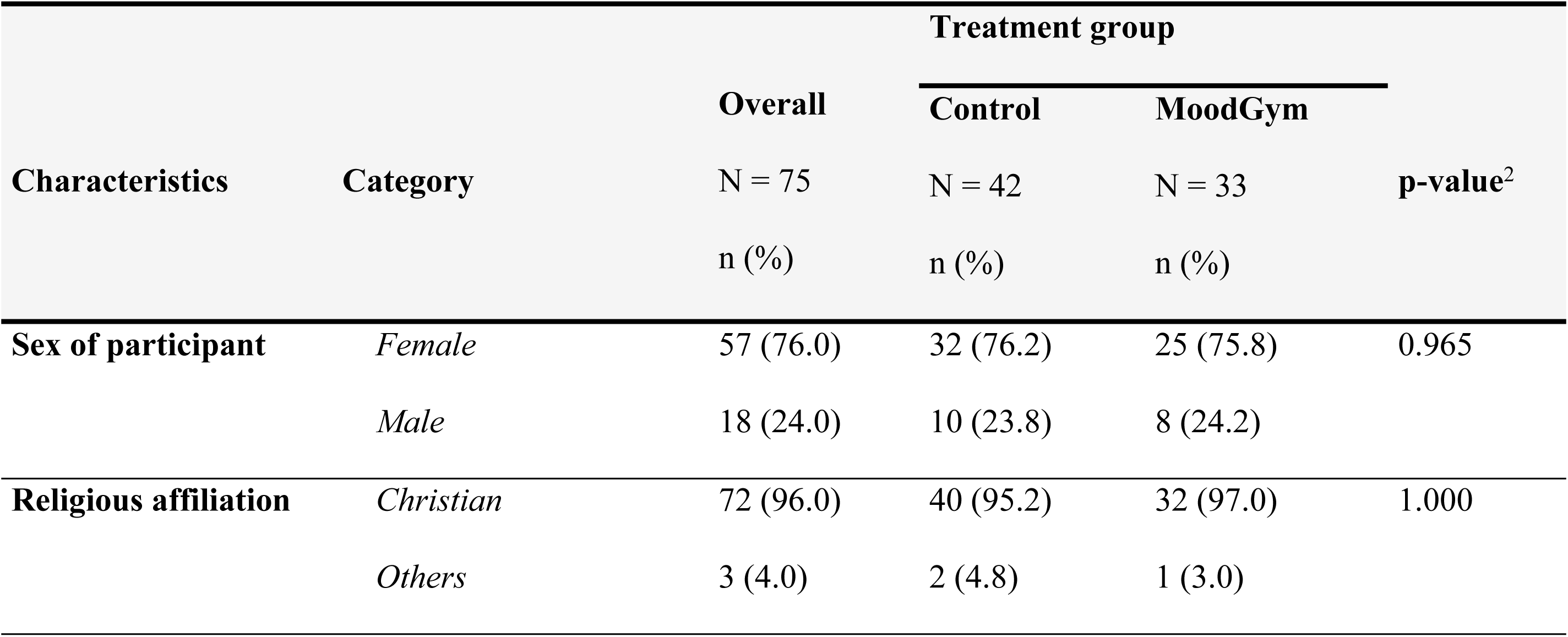
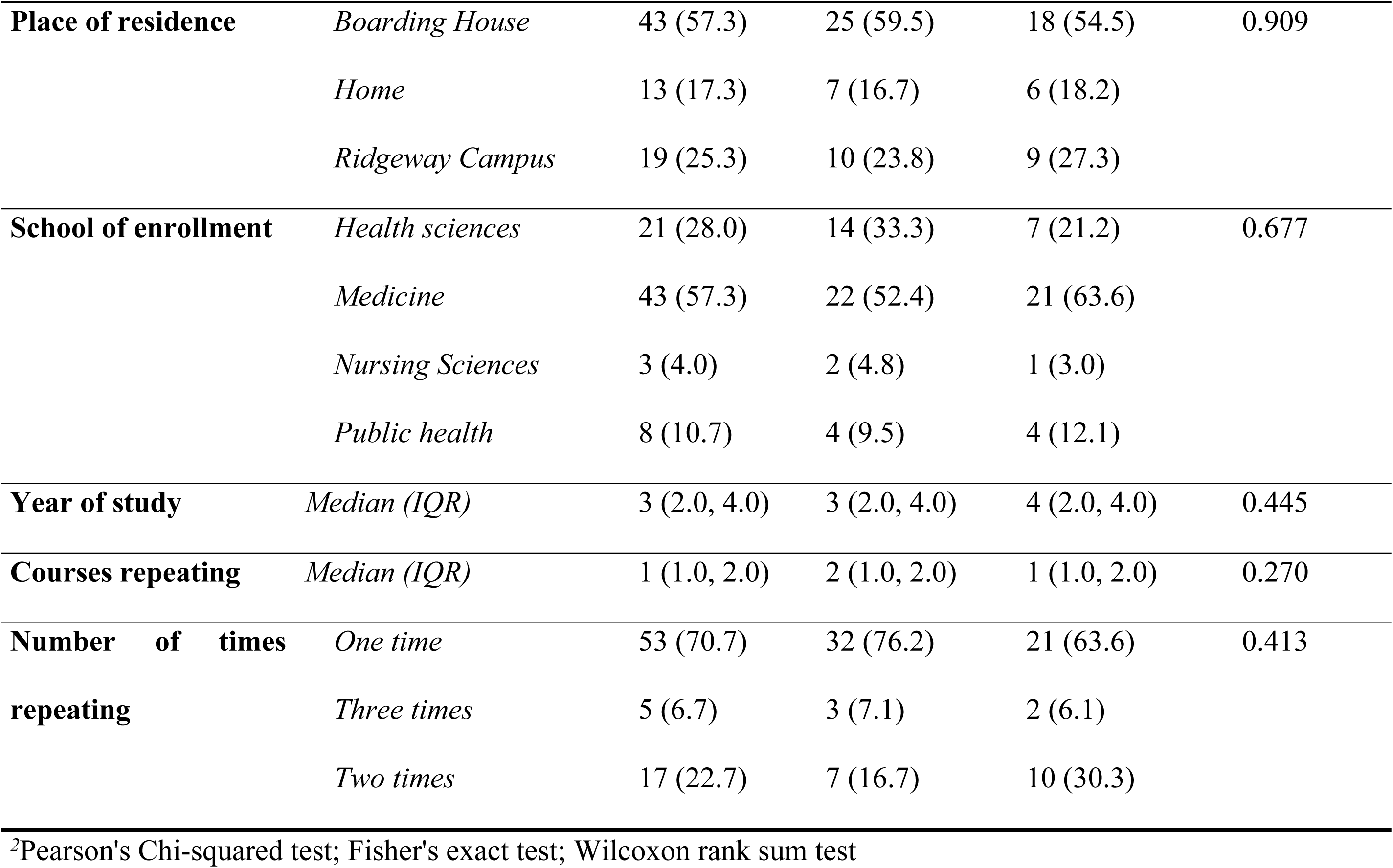
Comparison of baseline characteristics between treatment groups (n = 75)

The median year of study was 3 (IQR: 2–4), and the median number of courses being repeated was 1 (IQR: 1–2). Most participants (70.7%, n = 53) were repeating a course for the first time. There were no statistically significant differences between the intervention and control groups with respect to sex (p = 0.965), religious affiliation (p = 1.000), place of residence (p = 0.909), school of enrolment (p = 0.677), year of study (p = 0.445), number of courses repeated (p = 0.270), or number of times repeating (p = 0.413), indicating good baseline comparability between groups.

### Quantitative Findings

#### Baseline Depression Symptoms

Results in Table 4 show the baseline depression symptoms score and severity among study participants. Overall, participants had a mean score of 22 (±8.7), with the MoodGym group having a mean score of 24 (±9.7), while the control group had a mean score of 21 (±7.8). Both groups had comparable scores at baseline (p=0.196). At baseline, 18.7% (14) of the participants expressed symptoms of borderline clinical depression, 32% (24) had symptoms of moderate depression, and 14.7% (11) expressed symptoms of severe depression. About 3% (2.7%, 2) expressed symptoms of extreme depression. The distribution of depression severity levels was comparable between the treatment and control groups (p=0.147).

**Table 4:** Baseline levels of depression symptoms among study participants (n = 75)

|  | Treatment group |  |  | p-value<sup>2</sup> |
| --- | --- | --- | --- | --- |
|  | Overall | Control, | MoodGym, |  |
| Baseline symptoms | N = 75 | N = 42 | N = 33 |  |
|  | n (%) | n (%) | n (%) |  |
| Depression symptom score. Mean (±SD) | 22 (±8.7) | 21 (±7.8) | 24 (±9.7) | 0.196 |
| Severity of depression |  |  |  | 0.147 |
| Mild mood disturbance | 24 (32.0) | 16 (38.1) | 8 (24.2) |  |
| Borderline clinical depression | 14 (18.7) | 8 (19.0) | 6 (18.2) |  |
| Moderate depression | 24 (32.0) | 10 (23.8) | 14 (42.4) |  |
| Severe depression | 11 (14.7) | 8 (19.0) | 3 (9.1) |  |
| Extreme depression | 2 (2.7) | 0 (0.0) | 2 (6.1) |  |

<sup>2</sup>Two-Sample t-test; Fisher's exact test

#### Change in Depression Symptoms in the Intervention group

The distribution of depression symptom scores before and after participation in the MoodGym intervention among 33 participants is shown in Table 5. At baseline, the median depression score was 22 (IQR: 16–30), which decreased to 16 (IQR: 7–21) following completion of MoodGym. A Wilcoxon signed-rank test indicated that this reduction in depression symptom scores was statistically significant (Δ = 7, 95% CI: 4.5–9.5, p < 0.001). The Cohen’s *d* statistic demonstrated a large effect size for the MoodGym intervention in reducing depression symptoms in the intervention group (*d* = 1.02, 95% CI: 0.59–1.44).

**Table 5:**
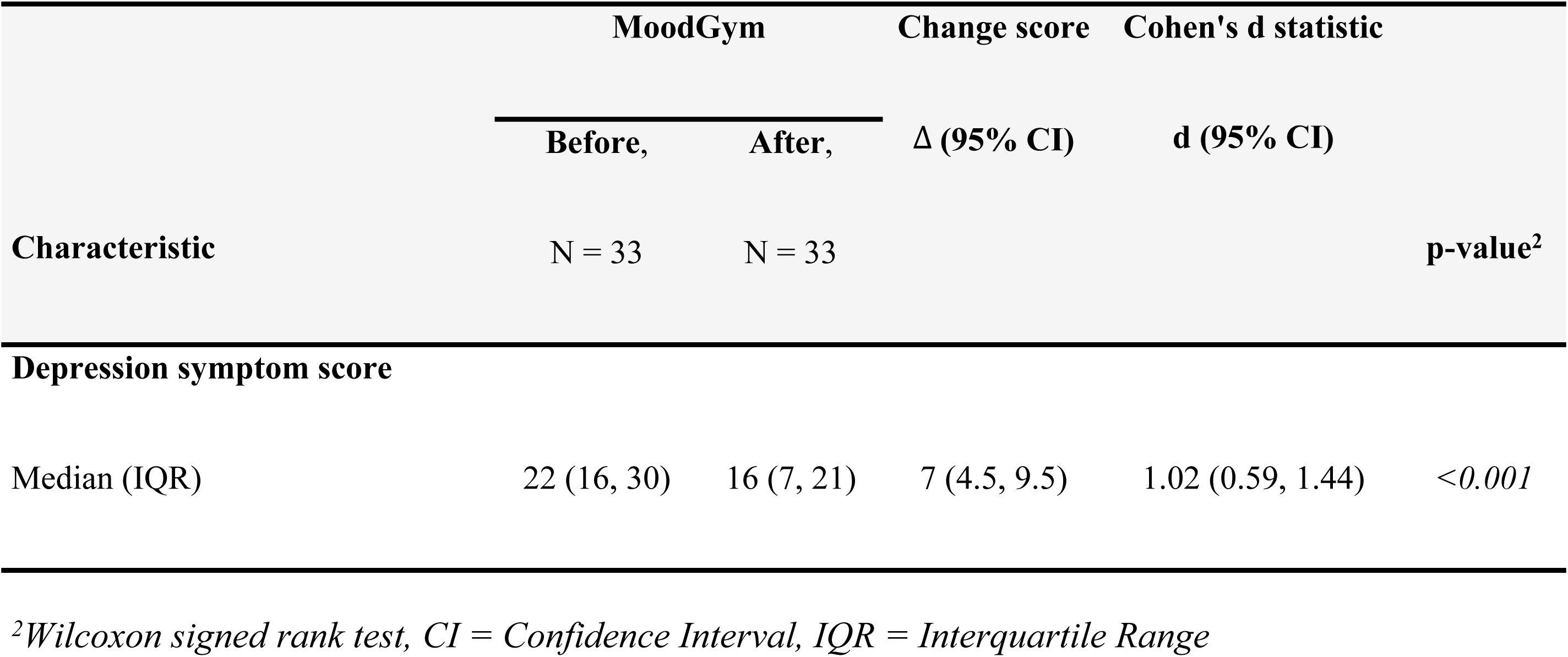
Depression symptoms score in the intervention group (n = 33)

|  | MoodGym |  | Change score | Cohen's d statistic |  |
| --- | --- | --- | --- | --- | --- |
| | Before, | After, | $\Delta$ (95% CI) | d (95% CI) | |
| Characteristic | N = 33 | N = 33 |  |  | p-value<sup>2</sup> |
| **Depression symptom score** |  |  |  |  |  |
| Median (IQR) | 22 (16, 30) | 16 (7, 21) | 7 (4.5, 9.5) | 1.02 (0.59, 1.44) | <0.001 |

<sup>2</sup>Wilcoxon signed rank test, CI = Confidence Interval, IQR = Interquartile Range

The change in median depression symptom scores over time among participants in the intervention group is illustrated in Figure 2. The figure shows that participants who received the intervention experienced a statistically significant reduction in depression symptoms from baseline to endline (22 vs. 16, p < 0.001).

**Fig 1:**
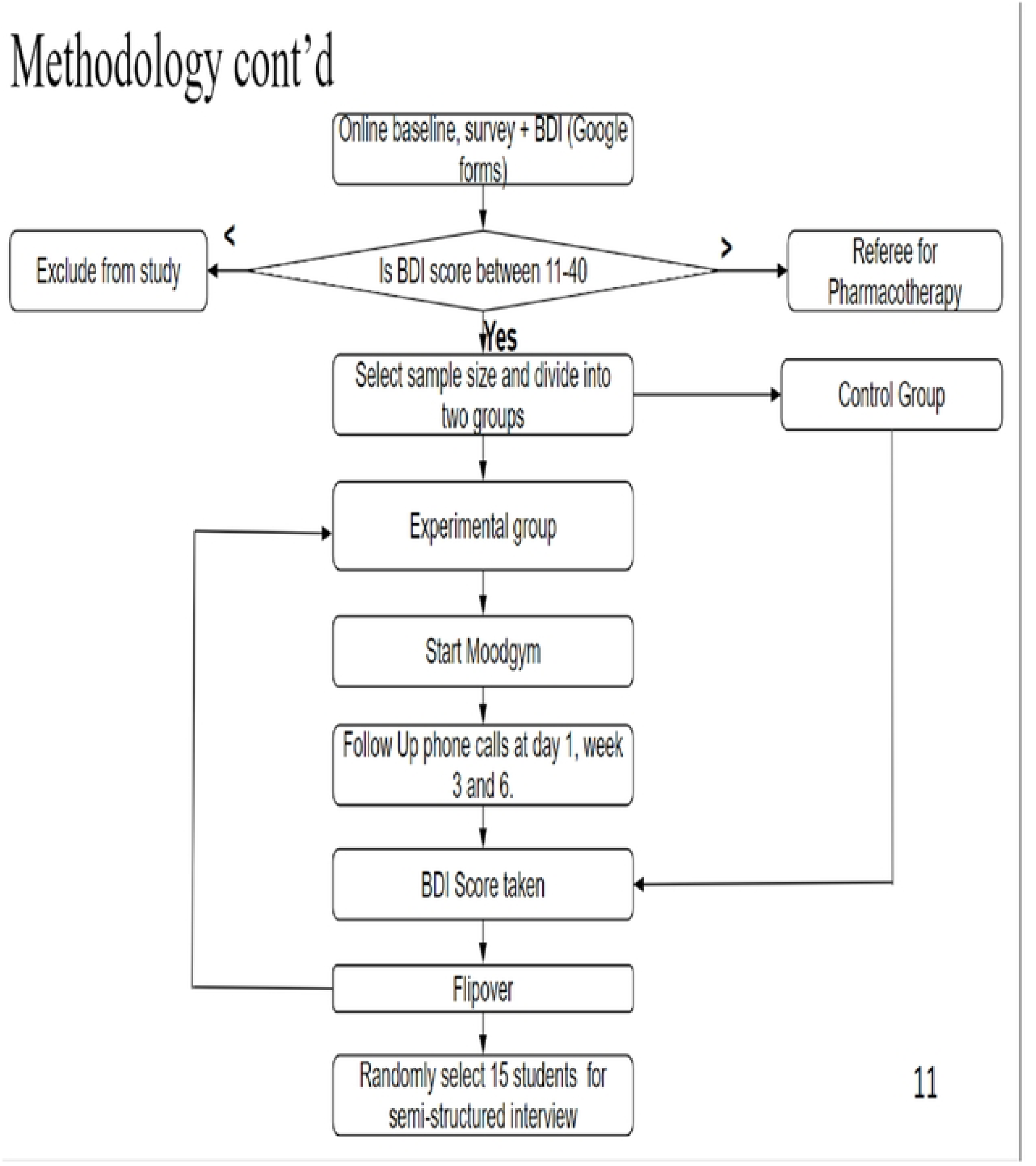
Flow of participants in the Study

**Fig 2:**
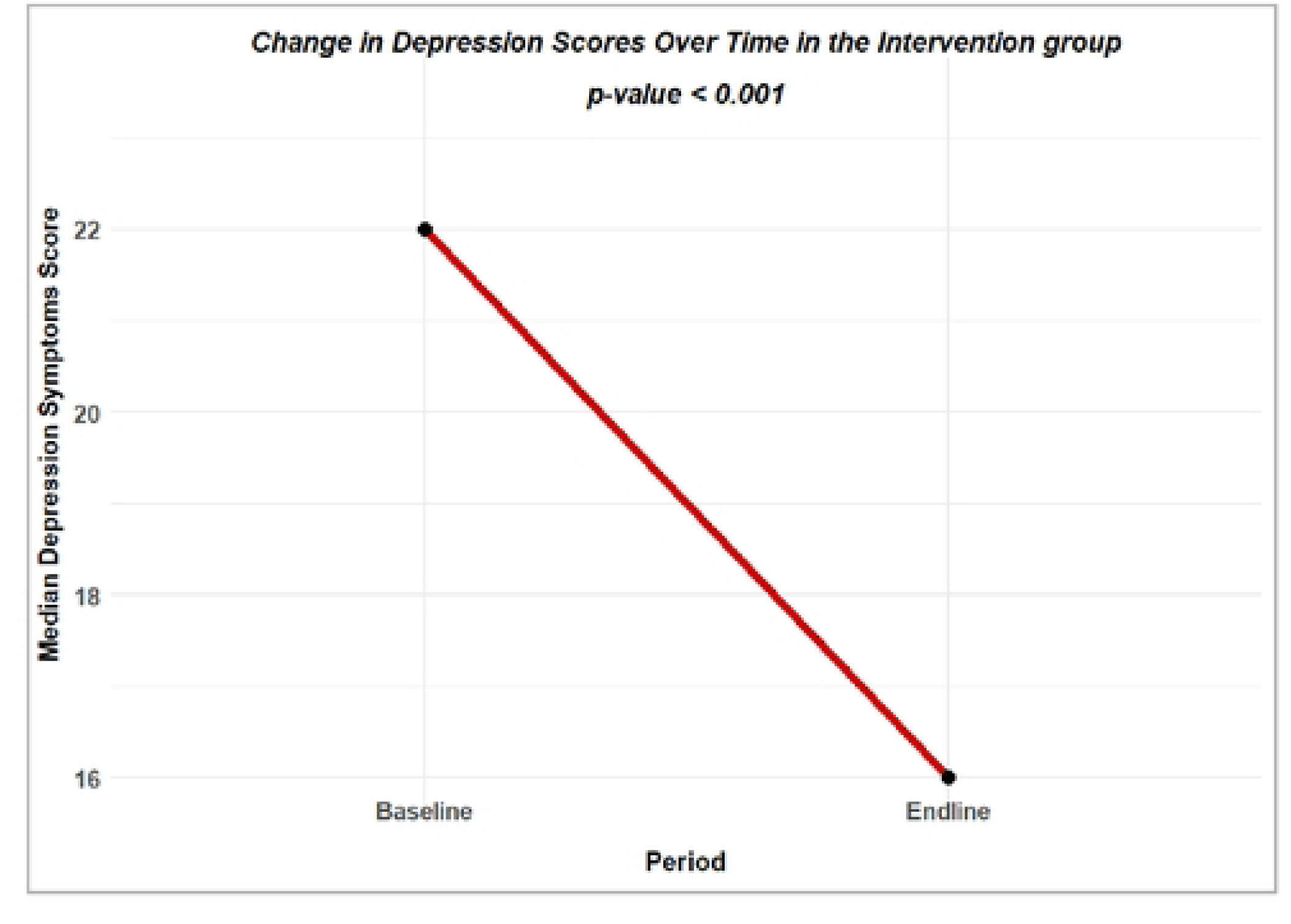
Change in depression symptom score in the intervention group (n = 33)

#### Change in Depression Symptoms between study groups

The difference-in-differences (DiD) analysis comparing changes in depression symptom scores between the MoodGym intervention group and the control group is shown in Table 6. The results indicate that participants in the MoodGym group experienced a 7-point reduction in depression symptom scores (IQR: −10 to −3), whereas the control group showed a median increase of 1.5 points between the baseline and endline periods (IQR: −2 to 10). The estimated difference-in-differences between the two groups was 9 points (95% CI: 7.0–12.0, p < 0.001), with the corresponding effect size indicating a large effect (*d* = 1.44, 95% CI: 0.92–1.94).

**Table 6:** Difference in differences of depression symptoms score between the control and intervention groups (n = 75)

| Characteristic | Treatment group |  | Difference in scores | Cohen's d statistic |  |
| --- | --- | --- | --- | --- | --- |
|  | Control, | MoodGym | Δ (95% CI) | d (95% CI) | p-value |
|  | N = 42 | N = 33 |  |  |  |
| Depression symptom score |  |  |  |  |  |
| Median change (IQR) | 1.5 (-2, 10) | -7 (-10, -3) | 9 (7.0, 12.0) | 1.44 (0.92, 1.94) | <0.001<sup>W</sup> |

*W* = Wilcoxon rank sum test, *CI* = Confidence Interval, *IQR* = Interquartile Range

The fixed-effects difference-in-differences model in Table 7 shows that depression symptom scores significantly reduced by about 10-point scores in MoodGym group over time compared to the control group (Coeff: −9.74, 95% CI: −12.80, −6.65, p<0.001). In contrast, depressive symptoms increased by over 2 points in the control group in the same time period (β = 2.17, p = 0.028). The intraclass correlation (ρ = 0.773) suggested that most of the variability in depression scores was due to between-individual differences, supporting the use of a fixed-effects approach.

**Table 7.** Fixed-effects difference-in-differences model estimating the effect of the MoodGym intervention on depression symptom scores (n = 75)

| Variable | Estimate ($\beta$) | Standard error | 95% CI | p-value |
| --- | --- | --- | --- | --- |
| Time (control group) | 2.17 | 0.96 | 0.11, 4.22 | 0.028 |
| MoodGym: Time | -9.74 | 1.60 | -12.80, -6.65 | <0.001 |
| rho (intraclass correlation) | 0.773 |  |  |  |

The trajectories of depression symptom scores from baseline to endline for the intervention (MoodGym) and control groups are illustrated in Figure 3. At baseline, both groups reported comparable depression scores (p > 0.05). Over time, the control group exhibited an increase in depression scores at endline, whereas the MoodGym group demonstrated a significant reduction in depressive symptoms across the study period (p < 0.001).

**Fig 3:**
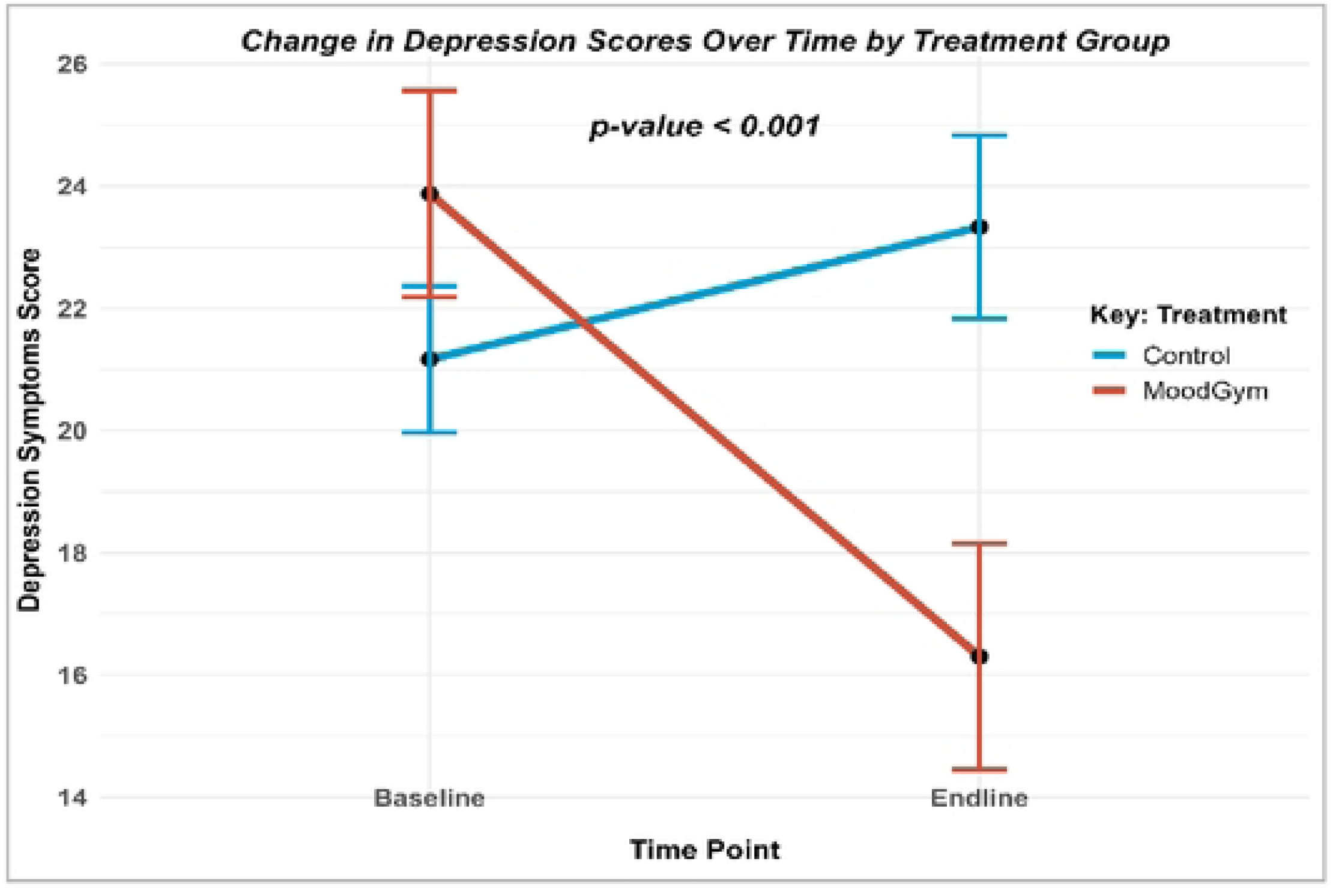
Difference in differences (DiD) analysis of the treatment effect of MoodGym on depression symptoms score (n = 75)

### Clinical Findings

This section presents the clinical interpretation of the quantitative study findings, focusing on changes in depressive symptom severity and clinically relevant outcomes associated with the MoodGYM intervention among repeating undergraduate students at the University of Zambia Ridgeway Campus. Clinical findings are interpreted using established severity thresholds of the Beck Depression Inventory (BDI) and are contextualised within student mental health, academic functioning, and neuropsychological well-being.

#### Baseline Clinical Profile of Depression

At baseline, the majority of participants presented with clinically significant depressive symptomatology, with a mean BDI score of 22 (±8.7), corresponding to moderate depression. More than two-thirds of participants fell within the borderline clinical, moderate, severe, or extreme depression categories, indicating that depressive symptoms extended beyond subclinical distress and were of potential clinical significance.

The presence of severe and extreme depressive symptoms in approximately 17% of participants is clinically concerning, given the established association between higher depression severity and impaired academic performance, reduced executive functioning, emotional dysregulation, and increased risk of suicidal ideation among medical and health sciences students. This baseline clinical profile underscores the heightened vulnerability of repeating undergraduate students, whose academic setbacks may intensify maladaptive cognitive patterns related to failure, hopelessness, and diminished self-efficacy—core features of depressive disorders.

Importantly, baseline comparability between the intervention and control groups across depression severity categories indicates that subsequent differences in clinical outcomes can be attributed with greater confidence to the MoodGYM intervention rather than pre-existing clinical differences.

### Clinically Meaningful Reduction in Depressive Symptoms Following MoodGYM

Clinically, participants who received the MoodGYM intervention demonstrated a substantial reduction in depressive symptom severity over the eight-week study period. The median BDI score decreased from 22 at baseline, indicative of moderate depression, to 16 at post-intervention, reflecting a shift toward mild depressive symptomatology.

From a clinical perspective, this reduction is both statistically and clinically meaningful, as it represents movement across established diagnostic severity thresholds. Such reductions are typically associated with improvements in functional capacity, emotional stability, and coping ability. The large observed effect size (Cohen’s d > 1.0) further supports the clinical relevance and therapeutic impact of the intervention.

In depression treatment research, a reduction of approximately 5–7 points on the BDI is widely considered indicative of a clinically meaningful response rather than normal symptom fluctuation. The magnitude of symptom reduction observed in this study therefore suggests that MoodGYM functioned as an active therapeutic intervention rather than solely a psychoeducational resource.

#### Worsening or Persistence of Symptoms in the Control Group

In contrast to the intervention group, participants in the control group exhibited a clinical trajectory characterised by persistence or worsening of depressive symptoms over the same study period, as reflected by an increase in median BDI scores. Clinically, this pattern is consistent with the natural course of untreated depression in high-stress academic environments, particularly among students experiencing repeated academic failure.

The divergence in symptom trajectories between the intervention and control groups highlights the protective effect of the MoodGYM intervention. Without structured psychological intervention, depressive symptoms in this population appear likely to persist or deteriorate, reinforcing the clinical importance of timely and accessible mental health interventions within university settings.

#### Functional and Psychological Clinical Implications

The observed reduction in depressive symptom severity among MoodGYM participants has important functional and psychological implications. Depression in university students is associated with impairments in attention, memory, executive functioning, and decision-making, all of which are critical for academic success in health-related disciplines. Clinically meaningful reductions in symptom severity are therefore likely to translate into improved cognitive efficiency, emotional regulation, and academic engagement.

From a neuropsychological standpoint, reductions in depressive symptoms may alleviate cognitive distortions, reduce psychomotor slowing, and enhance goal-directed behaviour. These changes are particularly relevant for repeating students, in whom depression-related cognitive impairment may further compromise academic recovery and progression.

#### Safety and Risk-Related Clinical Observations

From a clinical safety perspective, the study protocol appropriately excluded participants with extreme depression requiring immediate specialist or pharmacological intervention, referring such individuals to University Teaching Hospital services. No adverse clinical events or deterioration attributable to the MoodGYM intervention were observed during the study period.

The absence of symptom exacerbation in the intervention group supports the clinical safety and tolerability of MoodGYM as a low-intensity intervention for mild-to-moderate depressive symptoms. However, given the severity spectrum observed at baseline, the findings also reinforce the importance of clearly defined referral pathways for students with severe or complex depressive presentations.

#### Clinical Implications for Mental Health Service Delivery

The quantitative clinical findings support MoodGYM as a potentially effective intervention for reducing depressive symptoms among repeating undergraduate students in a low-resource university setting. Its demonstrated efficacy suggests that internet-based CBT may serve as a valuable component of stepped-care mental health models within university health services, particularly where access to specialist psychotherapy is limited.

The findings indicate that MoodGYM may be suitable as a first-line intervention for students with mild-to-moderate depression, with referral to higher-intensity clinical care for those with more severe symptomatology. Integration of such interventions into student mental health services could contribute to early symptom reduction, prevention of deterioration, and improved academic and psychological outcomes.

#### Summary of Clinical Findings

In summary, the quantitative clinical findings demonstrate that the MoodGYM intervention resulted in clinically meaningful reductions in depressive symptom severity among repeating undergraduate students, while participants who did not receive the intervention experienced persistence or worsening of symptoms. These results underscore the clinical importance of accessible, evidence-based psychological interventions for vulnerable student populations and provide a strong foundation for the discussion of broader implications in Chapter Five.

## Discussion

This chapter interprets the quantitative findings on the efficacy of MoodGYM, an internet-based cognitive behavioural therapy (iCBT) intervention, among repeating undergraduate students with depressive symptoms at the University of Zambia Ridgeway Campus. The discussion integrates (i) demographic and baseline clinical characteristics, (ii) quantitative treatment effects observed within and between study groups, and (iii) interpretation of findings in relation to existing CBT and iCBT literature.

### Demographic Characteristics

The participant profile reflects a student population in which academic strain and vulnerability to depressive symptoms are plausible, particularly in the context of course repetition. The baseline sample comprised 75 students, with females representing the majority (76.0%, n = 57) and males comprising 24.0% (n = 18). Religious affiliation was predominantly Christian (96.0%, n = 72), with a small minority reporting other affiliations (4.0%, n = 3). Regarding residence, more than half of the participants lived in boarding houses (57.3%, n = 43), while others lived at home (17.3%, n = 13) or on the Ridgeway Campus (25.3%, n = 19).

Academically, most participants were enrolled in the School of Medicine (57.3%, n = 43), followed by Health Sciences (28.0%, n = 21), Public Health (10.7%, n = 8), and Nursing Sciences (4.0%, n = 3). The median year of study was 3 (IQR 2–4), with a median of one repeated course (IQR 1–2). Most participants (70.7%, n = 53) were repeating a course for the first time. Importantly, treatment allocation resulted in comparable groups across measured baseline demographic and academic variables (all p values > .05), supporting the internal validity of subsequent outcome comparisons.

#### Interpretation and Implications

The demographic distribution—particularly the predominance of medical and health-sciences students—has important implications for baseline symptom burden and intervention relevance. Training in medical and health-related disciplines is characterised by high academic workload, frequent assessments, and performance pressure. For students who are repeating courses, these demands may be compounded by perceived academic failure, concerns about delayed progression, and heightened self-criticism, all of which are recognized risk factors for depression.

Residence patterns may also influence mental health outcomes and access to support. Students living in boarding houses or on campus may experience varying levels of privacy, social support, and environmental stressors, which can affect psychological well-being. These contextual factors underscore the importance of scalable interventions that can be accessed flexibly alongside academic commitments.

### Findings

#### Baseline Depression Symptom Burden

At baseline, participants demonstrated clinically meaningful levels of depressive symptoms. The overall mean depression score was 22 (SD ± 8.7), corresponding to moderate depression. Although the MoodGYM group had a slightly higher mean baseline score (24 ± 9.7) than the control group (21 ± 7.8), this difference was not statistically significant (p = 0.196), indicating adequate baseline comparability.

In terms of severity categories, 32.0% (n = 24) of participants were classified as having mild mood disturbance, 18.7% (n = 14) borderline clinical depression, 32.0% (n = 24) moderate depression, 14.7% (n = 11) severe depression, and 2.7% (n = 2) extreme depression. The distribution of depression severity did not differ significantly between the intervention and control groups (p = 0.147), strengthening confidence that subsequent outcome differences are attributable to the intervention rather than baseline imbalance.

These findings highlight the clinical relevance of addressing depression in repeating students. Depressive symptoms in academic contexts are associated with impaired attention, memory, motivation, and executive functioning, which are critical for academic recovery. This supports the rationale for evaluating accessible interventions such as iCBT programs that may reduce structural and psychological barriers to care (31).

#### Within-Group Pre–Post Changes in the MoodGYM Arm

A key quantitative finding was the statistically significant reduction in depressive symptom scores among students who received the MoodGYM intervention. In the intervention group (n = 33), the median depression score decreased from 22 (IQR 16–30) at baseline to 16 (IQR 7–21) post-intervention. This change was statistically significant based on the Wilcoxon signed-rank test (median change = 7; 95% CI 4.5–9.5; p < 0.001).

The effect size was large (Cohen’s d = 1.02, 95% CI 0.59–1.44), indicating a clinically meaningful magnitude of symptom improvement rather than a trivial or marginal effect. From a theoretical perspective, this pattern is consistent with the cognitive–behavioural model, which posits that modification of maladaptive cognitions and behaviours leads to reductions in depressive symptomatology (NHS, 2024).

#### Between-Group Effects: Difference-in-Differences and Fixed-Effects Modelling

Between-group analyses further strengthen the evidence for MoodGYM’s effectiveness. Difference-in-differences (DiD) analysis demonstrated that the MoodGYM group experienced a median reduction of 7 points in depression scores (IQR −10 to −3), whereas the control group showed a median increase of 1.5 points (IQR −2 to 10) over the same period. The estimated DiD effect was 9 points (95% CI 7.0–12.0, p < 0.001), with a large associated effect size (Cohen’s d = 1.44, 95% CI 0.92–1.94).

Consistent results were obtained from the fixed-effects DiD regression model, which estimated an approximate 9.74-point reduction in depression scores attributable to the MoodGYM intervention relative to controls (95% CI −12.80 to −6.65, p < 0.001). Over the same period, depressive symptoms in the control group increased by approximately 2.17 points (p = 0.028). The intraclass correlation coefficient (ρ = 0.773) indicates that a substantial proportion of variance was attributable to between-individual differences, supporting the appropriateness of the fixed-effects approach.

Interpretation

Taken together, the within-group and between-group quantitative findings indicate that MoodGYM produced a clinically and statistically meaningful reduction in depressive symptoms among repeating undergraduate students, whereas students who did not receive the intervention experienced persistence or worsening of symptoms. This pattern is consistent with existing evidence demonstrating the effectiveness of iCBT interventions in reducing depressive symptoms when CBT principles are delivered through structured online platforms (18,31).

## Conclusions

This study evaluated the efficacy of the internet-based cognitive behavioural therapy (iCBT) program MoodGYM in reducing depressive symptoms among repeating undergraduate students at the University of Zambia Ridgeway Campus. Consistent with the study hypothesis, the findings demonstrate that participation in MoodGYM was associated with statistically significant and clinically meaningful reductions in depressive symptom severity over an eight-week period, compared with no improvement—and in some cases worsening—among students who did not receive the intervention.

Students in the MoodGYM group experienced a large reduction in Beck Depression Inventory (BDI) scores, with effect sizes exceeding conventional thresholds for clinical relevance. The difference-in-differences and fixed-effects regression analyses further confirmed a robust treatment effect, with an approximately 9–10-point greater reduction in depression scores attributable to the intervention after controlling for baseline differences and time-invariant individual characteristics. In contrast, depressive symptoms in the control group persisted or increased over the same period, reflecting the likely natural course of untreated depression in a high-stress academic environment.

These results support the central hypothesis that structured, self-guided iCBT can effectively reduce depressive symptoms among academically vulnerable university students. The findings are consistent with prior evidence from high-income and middle-income settings demonstrating the efficacy of MoodGYM and other iCBT programs in student populations, and they extend this evidence base to a low-resource sub-Saharan African university context. Importantly, this study addresses a critical gap in the literature by focusing specifically on repeating undergraduate students, a group at elevated risk of depression that has been largely overlooked in previous digital mental-health research.

From a clinical and public-health perspective, the observed shift from moderate to mild depressive symptomatology among MoodGYM participants is particularly meaningful. Such reductions are likely to translate into improvements in emotional regulation, cognitive functioning, and academic engagement—domains that are essential for successful academic recovery among students repeating courses. The absence of adverse effects and the feasibility of delivering the intervention without intensive specialist input further support the suitability of MoodGYM as a low-intensity, first-line psychological intervention within stepped-care models of student mental-health service delivery.

The implications of these findings are substantial for universities in Zambia and similar low-resource settings, where access to face-to-face psychotherapy is constrained by shortages of trained mental-health professionals, stigma, cost, and time limitations. Internet-based CBT interventions such as MoodGYM offer a scalable, flexible, and potentially cost-effective approach to narrowing the mental-health treatment gap among university students. Integration of iCBT into campus health services, academic support programs, or student wellness platforms could facilitate early intervention and prevent the progression of depressive symptoms among at-risk students.

Despite its strengths, this study has limitations that warrant consideration. The quasi-experimental design limits causal inference relative to randomized controlled trials, and the sample size, while adequate for detecting meaningful effects, was smaller than initially planned. The study relied on self-reported depressive symptoms rather than diagnostic interviews, and longer-term outcomes beyond the eight-week intervention period were not assessed. Future research should therefore employ randomized designs, larger multi-institutional samples, and longer follow-up periods to evaluate the durability of treatment effects. Further work is also needed to examine engagement patterns, adherence predictors, cultural adaptation, and the impact of iCBT on academic outcomes such as course completion and progression.

In conclusion, this study provides strong evidence that MoodGYM is an effective and feasible intervention for reducing depressive symptoms among repeating undergraduate students in a low-resource university setting. The findings support the broader adoption of internet-based cognitive behavioural therapy as part of comprehensive student mental-health strategies and contribute important context-specific evidence to the growing global literature on digital mental health interventions.

## Data Availability

All relevant data are within the manuscript and its Supporting Information files.

## Acknowledgments

I wish to express my sincere gratitude to the University of Zambia for providing the institutional environment within which this research was conducted, and to the University of Zambia Biomedical Research Ethics Committee (UNZABREC) and the National Health Research Authority (NHRA) for their oversight, ethical approval, and regulatory guidance, which ensured that this study was conducted in accordance with national and institutional ethical standards. I am also deeply thankful to the research assistants and institutional stakeholders who provided invaluable support with participant recruitment, coordination, and data collection, and whose commitment greatly facilitated the successful implementation of the study. Special appreciation is extended to the statistician for expert guidance and support in data management and statistical analysis, which was essential to the rigour and validity of the study findings. Finally, I extend my heartfelt gratitude to all the students who voluntarily participated in this study and generously shared their time, experiences, and perspectives, without whom this research would not have been possible.

## Supporting information captions

**S1: Participant Consent Form**

**S2: Questionnaire**

